# Potential Community and Campus Covid-19 Outcomes Under University and College Reopening Scenarios

**DOI:** 10.1101/2020.08.29.20184366

**Authors:** James C. Benneyan, Christopher Gehrke, Iulian Ilieş, Nicol Nehls

**Affiliations:** Healthcare Systems Engineering Institute, Northeastern University, Boston, MA, USA

## Abstract

**Background:** Significant uncertainty exists in many countries about the safety of, and best strategies for, reopening college and university campuses until the Covid-19 pandemic is better controlled. Little also is known about the effects on-campus students may have on local higher-risk communities. We aimed to estimate potential community and campus Covid-19 exposures, infections, and mortality due to various university reopening and precaution plans under current ranges of assumptions and uncertainties.

**Methods:** We developed and calibrated campus-only, community-only, and campus-x-community epidemic differential equation and agent-based models. Input parameters for campus and surrounding communities were estimated via published and grey literature, scenario development, expert opinion, accuracy optimization algorithms, and Monte Carlo simulation; models were cross-validated against each other using February-June 2020 data from heterogeneous U.S. counties and states. Campus opening plans (spanning various fully open, hybrid, and fully virtual approaches) were identified from websites and publications. All scenarios were simulated assuming 16-week semesters and estimated ranges for Covid-19 prevalence among community residents and arriving students, precaution compliance, contact frequency, virus attack rates, and tracing and isolation effectiveness. Additional student and community exposures, infections, and mortality were estimated under each scenario, with 10% trimmed medians, standard deviations, and probability intervals computed to omit extreme outlier scenarios. Factorial analyses were conducted to identify intervention inputs with largest and smallest effects.

**Results:** As a base case with no precautions (or no compliance), predicted 16-week student infections and mortality under normal operations ranged significantly from 471 to 9,495 (median: 2,286, SD: 2,627) and 0 to 123 (median: 9, SD: 14) per 10,000 students, respectively. The maximum active exposures across a semester was 15.76% of all students warranting tracing. Total additional community exposures, infections, and mortality ranged from 1 to 187, 13 to 820, and 1 to 21 per 10,000 residents, respectively. 1% and 5% of on-campus students were infected after a mean (SD) of 11 (3) and 76 (17) days, respectively; >10% students infected by the end of a semester in 34.8% of scenarios, with the greatest increase (first inflection point) occurring on average on day 84 (SD: 10.2 days). Common reopening precautions reduced infections by 24% to 26% and mortality by 36% to 50% in both populations. Uncertainties in many factors, however, produced tremendous variability in all results, ranging from medians by −67% to +342%.

**Conclusions:** Consequences on community and student Covid-19 exposures, infections, and mortality of reopening physical campuses are very highly unpredictable, depending on a combination of random chance, controllable (e.g. physical layouts), and uncontrollable (e.g. human behavior) factors. Implications include needs for criteria to adapt campus operations mid-semester, methods to detect when necessary, and contingency plans for doing so.

## 1. Introduction

The Covid-19 disease pandemic has had devastating human, financial, and logistical impacts worldwide, including an estimated 19 million infected and 722,000 deaths^1^ (as of July 2020), radical changes to work and life routines, economic recession, and increased social inequities.^2-5^ Among many other issues, significant uncertainties exist about the safety, benefits, and potential consequences of reopening schools,^6-9^ heightened by resurgences in infections and mortality and campus-x-community cross-exposure concerns. ^9,10^ While much initial focus was on K-12 education, ^11-13^ similar college and university reopening concerns likely will continue for the next several academic terms. ^9,10,14^

As the Covid-19 pandemic spread uncontrollably during the spring of 2020, nearly all K-12 and secondary schools suspended physical classes, with an estimated 50 million elementary ^13^ and 19 million college U.S. students^15^ shifting to online learning, home schooling, and remote education, with experiences varied and often lacking. ^16-18^ While early in summer 2020 a few universities decided to remain fully virtual for the following academic year, ^19^ including the largest public university system in the United States, ^19,20^ many schools communicated their intent to re-open and started planning accordingly. Four events have since occurred of parallel import: several additional colleges and universities switched to full or partial on-line plans for the fall 2020 semester, Covid-19 has resurged in many regions, others have moved forward nonetheless to open as safely as possible, and debate has increased as to what best balances education, safety, and economic needs. ^21-26^

Examples of reopening plans range from full on-campus operations with contact precautions, hybrid virtual/physical formats with some courses (or class meetings within given courses) taught virtually and others in-person, having only first and/or second year students on campus with all others virtual, student choice to take any courses physically versus virtually, and (in the U.S.) accelerated semesters to end at Thanksgiving holiday to reduce travel-based spread. ^27-29^ Efforts to limit on-campus exposures include reconfigured classrooms and dormitory spaces, precaution awareness campaigns, hotel room rentals to reduce living density, testing and tracing plans of varied rigor, returning student isolation, dedicated living spaces tested-positive students, and other strategies that attempt to reduce density and exposure rates. ^21,23,27,30-32^

Significant uncertainty, however, exists about the effectiveness of any of these plans. ^21,33,23,31^ The best current diagnostic tests have variable and poor clinical sensitivity, ^25,34^ an estimated 30-40% of positive individuals never exhibit symptoms, ^25,34,35^ incubation delays from exposure to symptomatic average 3-5 days, ^35-37^ and on-campus compliance to distancing precautions. ^25,30,38,39^ Contact tracing, while helpful, may not work as well for Covid-19 given the above ^9,33^ and may be further limited here as students interact with many-fold more individuals (many unknowingly or unknown by name).

These uncertainties have prompted some to question university reopening safety, ^6,8,14,26,31,40^ ^25^ especially in urban university settings with significant geographically dispersed student populations.^41^ Others have suggested Covid-19 might catalyze reinvention of higher education, ^42-45^ including criticisms of prioritizing economics, brand, and survival over safety. ^22,43,44,46^ The president of Paul Quinn College, for example, recently stated “Rushing to reopen our society and our schools is a mistake that will ultimately result in hundreds of thousands of citizens falling sick and worse. We should not let our own financial and reputational worries cloud our judgment about matters of life and death.” ^8^ As elsewhere, not reopening may have large economic and student development effects, ^47-49^ although perhaps less so the latter than K-12 schools. Not reopening could be untenable for colleges and universities already facing financial strains before Covid-19 emerged. ^48-50^

Although little empirical data exist yet on college reopening, ^40,51-56^ experiences of pre-school, summer camp, and K-12 programs have been varied, ^57,58^ with some outbreaks traced back to only a few index cases. ^21^ Social gatherings of college-age students during summer 2020 also have resulted in outbreaks, ^38,58,59^ including events and activities individuals advised against but participated in nonetheless. ^30,38^ Despite early uncertainty, increasing evidence suggests student-aged individuals can carry and transmit the SARS-CoV-2 virus ^35,58,60,61^ and significant between-student spread occurs at college and high school levels ^35,58,61^ (in contrast to younger K-5 students). ^35,47,61,62^ The impacts of campus opening on spread to the surrounding community, with higher percentages of at-risk individuals, has been less reported on.

Given these combined uncertainties, we developed and applied single and multiple population Covid-19 spread models to investigate the range of potential community and campus impacts across a range of potential reopening scenarios. The intent is to provide model-based analysis to better inform, among other inputs, decision-making at a critical time in the Covid-19 pandemic. While similar model analyses have extensively studied other infectious disease policies, ^63-66^ little investigation has focused on university reopening.

## Methods

### Model Development and Validation

We developed and validated single and multiple population differential equation (ODE) and agent-based models of Covid-19 spread within and between defined groups of individuals. The general model logic (Figure 1) was adapted from classic susceptible-exposed-infected-recovered (SEIR) frameworks ^67,68^ similar to those reported on elsewhere for many other infectious disease concerns. ^63-76^ The single population model describes spread dynamics within one defined population (e.g. on-campus students or local community residents) depending on input values, whereas the multi-population model additionally includes cross-exposure between two or more groups. All models were adapted from validated MatLab code used to study other epidemics over several years.

**Figure 1.**
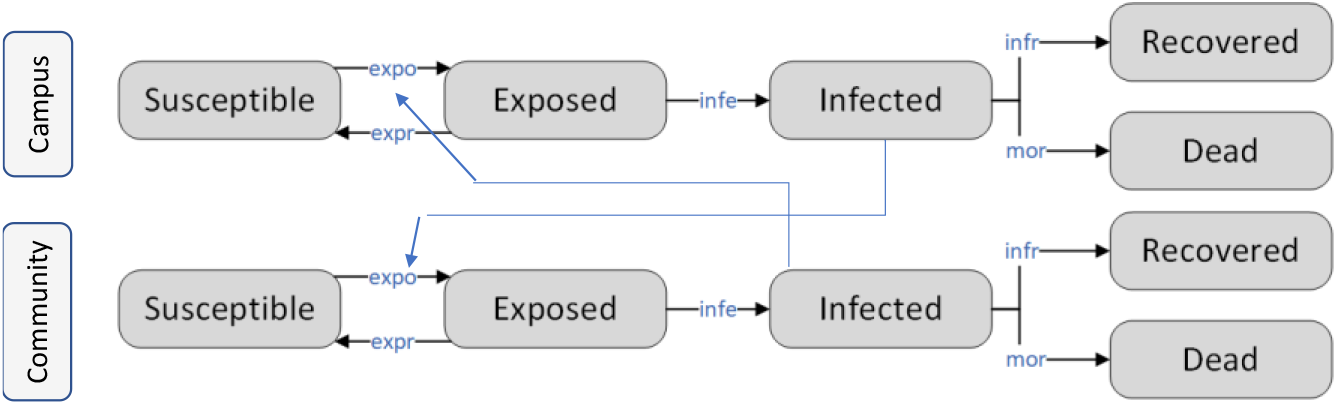
General logic of campus-x-community two-population Covid-19 disease spread model. (Suspectible: Individuals not currently infected but who can become infected; Exposed: Individuals who are exposed and potentially infected but not infectious yet to others; Infected: Individuals who are infected and can infect others; Recovered: Individuals who were infected, survived, and cannot become re-infected nor infect others within the study timeframe; Dead: Individuals who were infected and died from Covid-19 or complications).

State variables at time *t* include the numbers of individuals in population *j* that are Covid-free and susceptible (*S_j_*(*t*)), exposed to Covid but not yet infectious themselves (*E_j_*(*t*)), Covid-positive and infectious to others (*I_j_*(*t*)), recovered and not susceptible to re-infection (*R_j_*(*t*)), and Covid-associated deceased (*D_j_*(*t*)). Recovered individuals are assumed not to be able to re-infect, at least not with any appreciable rate within a one semester timeframe. ^37,77^ Multiple dynamics change points were included for all parameters to allow for policy or behavior changes when fitting models to historical data. Each state variable is updated numerically at each time increment based on its previous value, current values of other state variables, and equations governing their interdependent relationships, with this process continuing iteratively for 16 weeks.

For example, the number of individuals in the susceptible population (*S_j_*(*t*)) is decremented by the number of newly exposed individuals (*S_j_*(*t*) · *expo_j_*) and increased by the number who previously were exposed but did not develop infections (*E_j_*(*t*) · *expr_j_*), where the daily exposure rate *expo_j_*, the average risk of transmission multiplied by the average number of contacts per day, is back-computed from the basic reproduction number *R*_0_ (average number of new infections per infected individual) and recovery and mortality rates, and the recovery rate of non-infectious exposed individuals *expr_j_* is calculated as the inverse of the corresponding recovery time *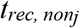*.

In turn, the number of exposed individuals is increased by *S_j_*(*t*) · *expo_j_* and decremented by the number who develop infections (*E_j_*(*t*) · *infe_j_*), where the daily infection rate *expo_j_* is the ratio of the probability of becoming infected upon exposure *p_j_* over the average incubation time 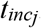. Infected individuals either recover or die at rates of *infr_j_* · *I_j_*(*t*) and *mort_j_* · *I_j_*(*t*), respectively, where the daily recovery and mortality rates are the inverse of the average recovery time *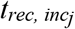* and the ratio of the overall COVID case fatality rate for that population (*CFR_j_*) over the average time from infection until death *t_i2dj_*, respectively. The change in each state variable and their governing rate change dynamics at each time step during numeric evaluation thus are:

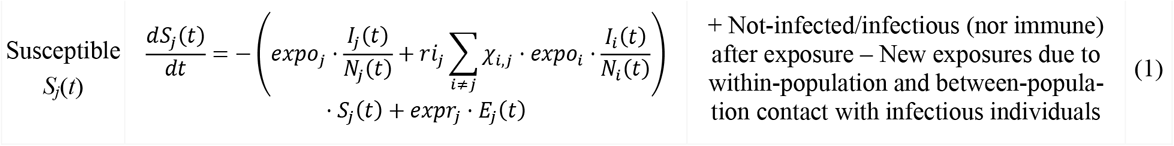

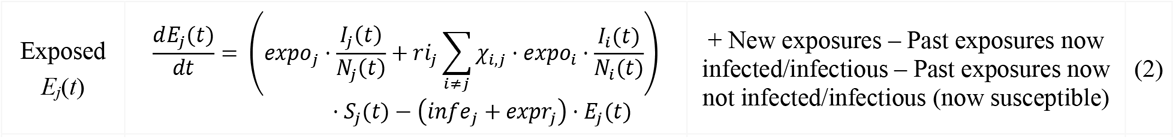

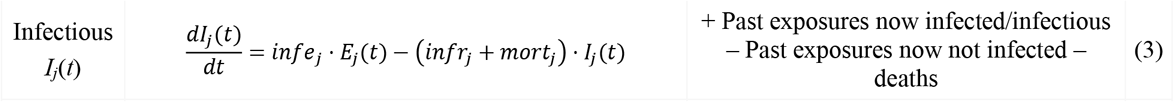

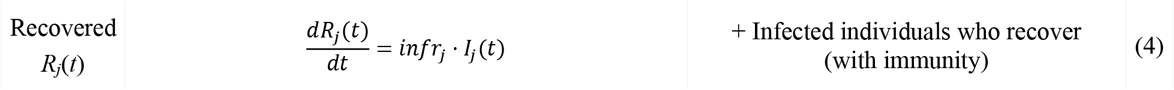

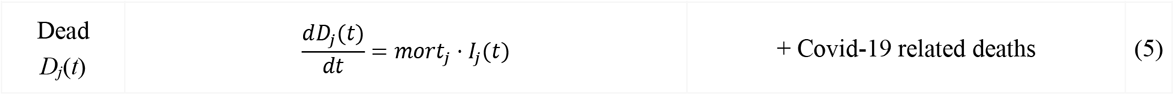

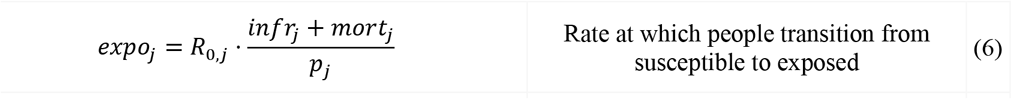

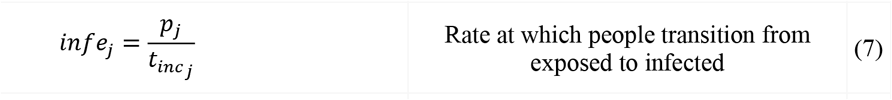

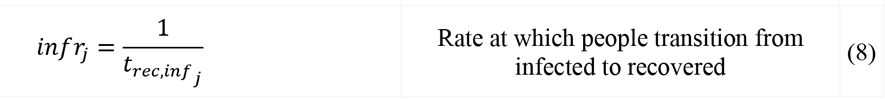

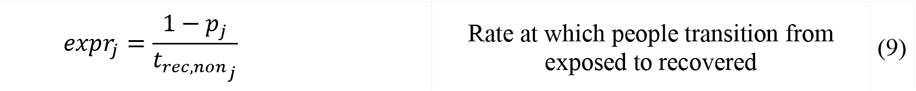

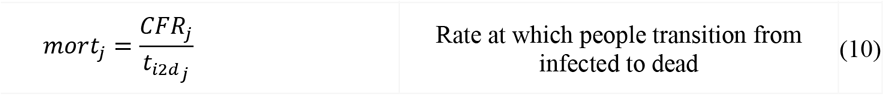

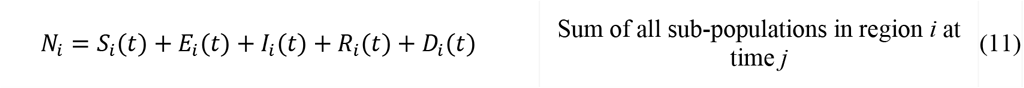

where *χ_i,j_* = 1 if populations *i* and *j* are connected and 0 otherwise and *p_j_* = the proportion of exposed individuals that transition to infected (versus recovering to susceptible). The multi-population models allow for separate parameter values for each population, such as based on their demographics, with a cross-exposure parameter (*ri_j_*) defining the relative rate at which infectious individuals in one population expose susceptible individuals in the other (typically lower than within-population, assuming less interaction).

### Parameter Estimation and Calibration

Model accuracy was validated using standard methods, ^65,78-82^ cross-validation, and varied state and county empirical data (Jan-July 2020) exhibiting different epidemic patterns, magnitudes, and timings (Figure 2). In all cases model results closely emulate historical data, with accuracy on par with or exceeding norms and results reported elsewhere ^83-89^ and with ≤ 1 change points generally providing good fits, suggesting good prospective short-term prediction capability.

**Figure 2.**
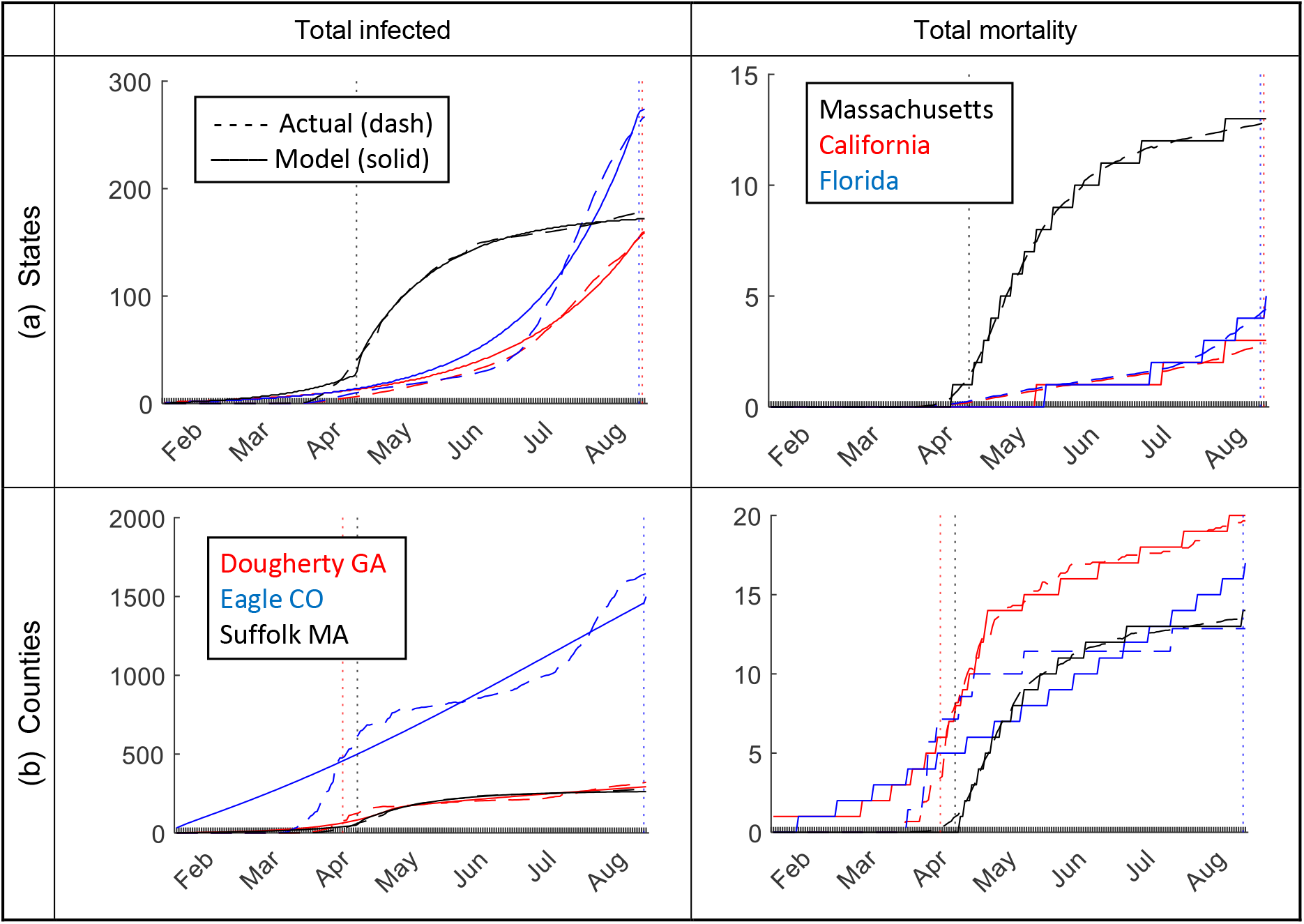
Examples of model accuracy; (a) U.S. state (shown: Massachusetts, California, and Florida) and (b) county (shown: Dougherty county GA, Eagle county CO, Suffolk county MA).

Table 1 summarizes input estimates used in the community and campus models, estimated as described above. For inputs with uncertainty, we used Monte Carlo simulations to create 1,000 synthetic results across plausible ranges, using the shown most likely, maximum, and minimum values to generate asymmetrical triangular distribution random variates. Since little data yet exist about on-campus spread, ^21^ for exposure rates we used the shown ranges for the average number an infected student infects divided by the exposed-to-infectious percentage.

For community populations, we further calibrated inputs via meta-heuristic search algorithms (particle swarm, genetic algorithms, simulated annealing) to minimize root mean square error differences between historical and model-predicted infections and mortality, running each parameterization 1,000 times to produce 1,000 optimized input sets. For model fits with change points, separate values for all inputs were optimized for each time segment, with state variables at the start of each new time segment set to their values at the end of the prior segment.

For initial disease prevalence in the local community and among arriving students, we also used expected values and probability intervals from a validated epidemic data curve fitting algorithm. In the latter case, we predicted Covid-19 community prevalence at the start of the semester among arriving students using a proportionally weighted average of prevalence predictions based on home locations. Resulting community and student prevalence ranges were validated against data reported in recent media. Positive individuals at semester start-up were assumed distributed between exposed but not yet infectious (24.3%) and infectious (75.7%) groups based on approximate relative durations an average infected individual might spend in each state. For all analyses, 10% trimmed medians, standard deviations, and percentile intervals were computed to reduce any extreme outlier scenario effects.

**Table 1.** Model parameters in Covid-19 campus-x-community epidemic models, estimated values, and ranges used for parameter search and sensitivity analysis. “Rank order” indicates relative significance of each parameter on campus (community) outcomes (16 week totals); only statistically significant factors shown (*α* = .05).

| Parameter | Definition | Lower Bound | Most Likely | Upper Bound | Sources | Rank |  |
| --- | --- | --- | --- | --- | --- | --- | --- |
|  |  |  |  |  |  | Infection | Mortality |
| $R_{0,1}$ | Average number of students who become infected by infectious students | 0.66 | 1 | 3.4 | [90] | 2 (6) | 5 (—) |
| $R_{0,2}$ | Average number of residents who become infected by infectious residents | 0.66 | - | 3.4 | [90], parameter search | 5 (2) | — (5) |
| $ri_j$ | Cross-exposure parameter (campus x community) | 0.005 | 0.008 | 0.02 | estimated | 6 (6) | — (—) |
| $\pi_1$ | Proportion of student population that is initially infected at semester start | 0.001 | .01 | .05 | [91-93] | 4 (—) | 6 (—) |
| $\pi_2$ | Proportion of community population that is initially infected at semester start | 0.0016 | .01 | .016 | [94] | — (4) | (7) |
| $p_j$ | Proportion of exposed people that become infected | 0.5 | 0.9 | 1 | estimated | — (7) | 3 (3) |
| $t_{inc j}$ | Incubation duration (in days) | 2 | 4.5 | 14 | [95,96] | 3 (3) | 7 (6) |
| $t_{rec j}$ | Recovery duration (days) | 6 | 14 | 42 | [97-99] | 1 (1) | 2 (1) |
| $CFR_1$ | Fatality rate for college population | 0.001 | 0.0092 | 0.016 | [97,100] | — (—) | 1 (—) |
| $CFR_2$ | Fatality rate for community population | 0.01 | 0.06 | 0.15 | [101,102] | — (5) | — (2) |
| $t_{i2d j}$ | Number of days from infection until death | 14 | 35 | 56 | [99] | — (8) | 4 (4) |

### Reopening Scenario Analysis

Common university reopening scenarios were identified from literature and published surveys, ^19^ generally belonging to one of several categories (Table 2); a recent New York Times survey (July 29, 2020) ^7^ also summarized reopening plans of 271 U.S. colleges and universities. The most common approaches included primarily or fully in-person (35%), primarily or fully online (32%), and hybrid (19%). For example, the University of Washington reopening plan ^103^ illustrates a common approach wherein more than 90% of courses will be taught online, only with courses that cannot be taught remotely (e.g. medical and health sciences) taught in person with safety precautions; the majority of student services and advising will take place remotely, and any staff who can work remotely will continue to do so. In contrast, Purdue University illustrates an opposite approach ^104^ wherein classes mainly will be taught on campus with contact precautions until Thanksgiving break and relying on students to manage their personal safety.

**Table 2.** Representative examples of university and college Covid-19 fall 2020 semester physical reopening plans.

| Intervention Description | Examples | Source | Estimated Reduction in $R_0$ |
| --- | --- | --- | --- |
| Remote coursework for classes over 50, testing, contact tracing, health surveillance in dorms | University of Washington | [103, 105] | 36% |
| Remote option available, social distancing, shortened semester, flexible start dates for international students | Rice University | [106] | 25% |
| Face masks, social distancing, limited classes, some coursework online, fewer students living on campus, shorter semester | Stanford University | [107] | 33% |
| More online classes, masks, social distancing, testing, health surveillance, sanitizing and washing stations | Ohio State University | [108, 109] | 30% |
| Some classes online, expanded housing, social distancing, face masks, staggered hours, increased cleaning, testing, tracing | Northeastern University | [32, 110] | 49% |
| Most classes online except those for which in-person instruction is deemed necessary | California State Universities | [111, 112] | 12% |
| Students back on campus for a shortened fall semester as long as they follow CT reopening suggestions | Connecticut State Institutions | [113, 114] | 46% |

For each scenario, 1,000 model replications were run for campus alone, community alone, and campus-x-community together to estimate additional cross-exposure impacts of each population on the other. For the campus-x-community cases, each of the 1,000 community parameterizations were randomly coupled with the 1,000 random sets of campus inputs, with the two populations interacting using 1,000 random values of the cross-exposure parameter, *ri_j_*, sampled from the range shown in Table 1. Overall results (medians, standard deviations (SD), and 95% probability intervals) and pairwise differences were computed for and between the 1,000 campus results and their campus-x-community counterparts, and similarly the 1,000 community results and their campus-x-community counterparts, in order to estimate attributable cross-exposure effects on community residents and students.

We assumed three geographic settings: (1) an urban campus of 10,000 students with 100,000 residents living the immediately surrounding residential areas or neighborhoods in which off-campus students tend to reside, (2) the same size student body (10,000) but now with fewer (40,000) residents living near campus, and (3) a smaller number of 2,000 students with 40,000 residents living near campus. The first scenario might represent a large university in a major city, whereas the second might represent a large rural university and the third a smaller undergraduate college in a non-urban setting. These student-to-community populations (1:10, 1:4, 1:20) can be extrapolated to other settings with similar ratios.

All results were tabulated, plotted longitudinally, and verified independently. First inflection points (dates of steepest increases) for each outcome, scenario, and population were identified numerically since in diffusion theory interventions after these points tend to be less effective. Sensitivity analyses were conducted to identify inputs to which results are most sensitive (via central composite factorial experimental designs ^115^) as this could inform policy-making, interventions, and target setting.

## Results

### On-Campus / Student Impact

Depending on Covid-19 prevalence among arriving students and semester initialization precautions, Figure 3 summarizes the predicted number of students per 10,000 who could be exposed, infectious, and die over. By semester end, under the base case (2% arrival prevalence, little returning precautions and/or effectiveness) student outcomes may range from 471 to 9,458 infections (median: 2,286, SD: 2,627) and 0 to 123 deaths (median: 9, SD: 14). The more realistic case (1% prevalence after intake) reduces these consequences to a median (SD) of 1,332 (2,552) infections and 5 (12) deaths, with the greatest exposure and infection increases typically occurring at mid-semester onwards (with important implications on non-local spread as students return to their home communities).

**Figure 3.**
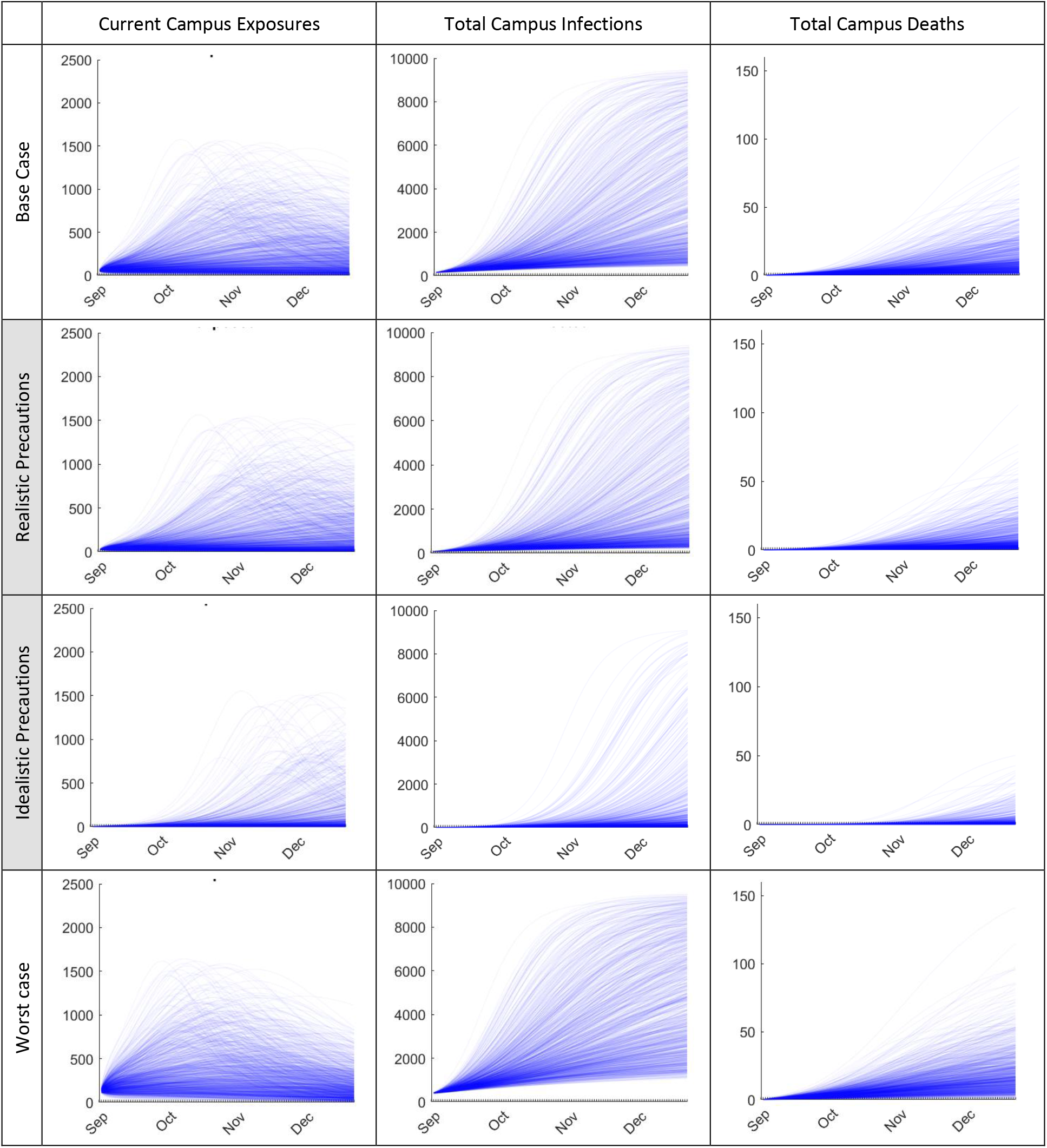
Predicted number of students per 10,000 who are currently exposed, total infected, and total mortality over a 16-week semester. Top row: base case scenario assuming no semester initiation precautions and disease prevalence of 2% among arriving students equal to national and regional averages. Shaded middle rows (most likely cases): realistic (1%) and idealistic (0.1%) initial prevalence scenarios assuming good or great screening-on-arrival precautions, adherence, and effectiveness. Bottom row: worst case scenario (5%) assuming home state rate increases and little-to-no arrival precautions, compliance, or effectiveness.

While less likely, the idealistic (0.1%) and worst-case (5%) scenarios were included for reference comparison. The first would result in a median (SD) of 158 (1,760) infections and 1 (6) deaths, and the latter 3,996 (2,485) infections and 16 (17) deaths. Under the base case, the total number of active student exposures that under a contact tracing approach should be identified range from 3 to 1,576 with implications on resource planning and viability. Under the two most likely scenarios (Table 3) by mid-semester consequences might reach as high (mean plus one standard deviation) as 810 infections and 4 deaths per 10,000 students, or conversely as low (mean minus one standard deviation) as 782 infections and 0 deaths.

**Table 3.** Predicted median number of monthly exposures, infections, and mortality per 10,000 students. Expo: exposures; Infect: infections; Mort: mortality. Values in parentheses indicate 95% probability ranges given uncertainties in many assumptions and inputs.

| scenario | September |  |  | October |  |  | November |  |  | December |  |  |
| --- | --- | --- | --- | --- | --- | --- | --- | --- | --- | --- | --- | --- |
|  | Expo | Infect | Mort | Expo | Infect | Mort | Expo | Infect | Mort | Expo | Infect | Mort |
| Base Case | 474<br>(175, 1840) | 318<br>(146, 1181) | 1<br>(0, 3) | 708<br>(160, 4731) | 510<br>(131, 3561) | 2<br>(1, 11) | 925<br>(124, 3532) | 685<br>(105, 2946) | 3<br>(0, 16) | 638<br>(74, 1934) | 522<br>(63, 1576) | 2<br>(0, 12) |
| Realistic precautions | 245<br>(89, 1011) | 163<br>(74, 639) | 1<br>(0, 2) | 392<br>(84, 3723) | 278<br>(68, 2777) | 1<br>(0, 8) | 554<br>(67, 4024) | 411<br>(56, 3304) | 2<br>(0, 14) | 504<br>(41, 2279) | 389<br>(36, 1874) | 2<br>(0, 12) |
| Idealistic precautions | 26<br>(9, 113) | 17<br>(7, 69) | 0<br>(0, 0) | 44<br>(9, 769) | 30<br>(7, 476) | 0<br>(0, 1) | 70<br>(7, 3072) | 49<br>(6, 2133) | 0<br>(0, 5) | 76<br>(5, 2997) | 56<br>(4, 2366) | 0<br>(0, 8) |
| Worst case | 1084<br>(415, 3613) | 744<br>(353, 2398) | 3<br>(1, 8) | 1377<br>(354, 4623) | 1026<br>(301, 3895) | 5<br>(1, 17) | 1225<br>(248, 2840) | 994<br>(221, 2380) | 5<br>(1, 19) | 624<br>(122, 1406) | 537<br>(114, 1214) | 3<br>(1, 12) |

Figure 4 summarizes similar results assuming various semester-long operation precautions, with 25%-50% reductions plausible from effective testing, tracing, and isolation. In general, strategies to reduce exposure are very effective, as is intuitive, although under most scenarios a concerning number of students still can become infected or die. For example, in the very optimistic case with a 75% reduction in close contact, 95 to 132 infections (median: 107) and 1 death per 10,000 students may be likely by mid-semester, increasing to 97 to 139 infections and 0 to 3 deaths by semester end.

**Figure 4.**
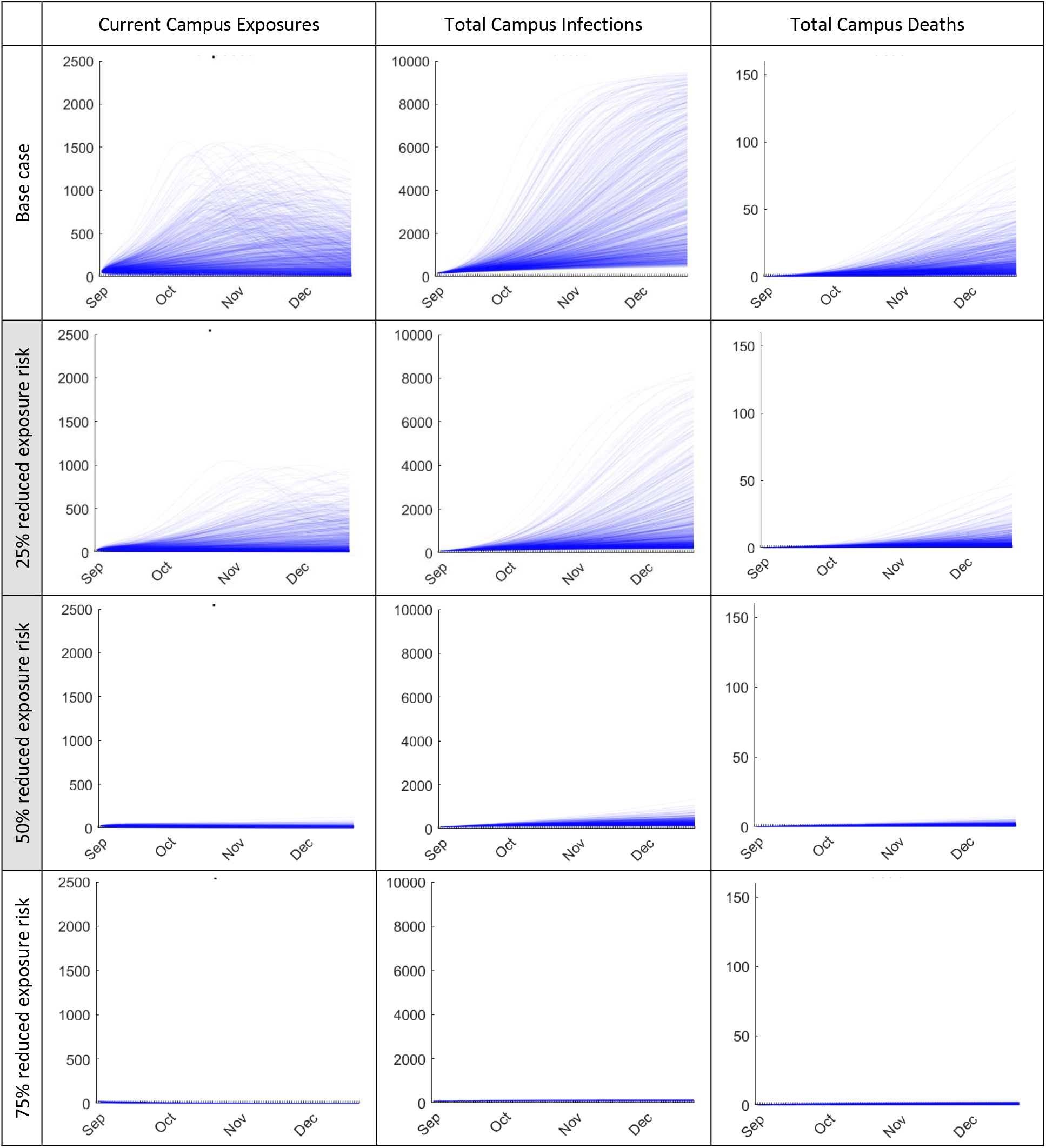
Relative effectiveness of reopening and precaution strategies on reducing campus exposures, infections, and mortality per 10,000 students (assuming 1% of students are infected or exposed at the start of the semester). Top row: base case from Figure 3 for comparison; Shaded middle rows (most likely cases): realistic precaution effectiveness and compliance cases; Bottom row: idealistic precaution effectiveness and compliance. (Reduced exposure risk refers to reducing *R_0_*.)

### Community Resident Impact

Figure 5 summarizes additional community (blue lines) and campus (red lines) impacts of reopening due to campus-x-community cross-exposure, assuming the same scenarios described above; for comparison the top row shows the baseline number of community exposures, infections, and mortality without reopening. The local community impacts (Table 4) of opening with little-to-no semester operation precautions and/or adherence might range from 1 to 9,768 additional community infections (median: 158, SD: 1,131) and 0 to 491 additional community deaths (median: 6, SD: 53).

**Figure 5.**
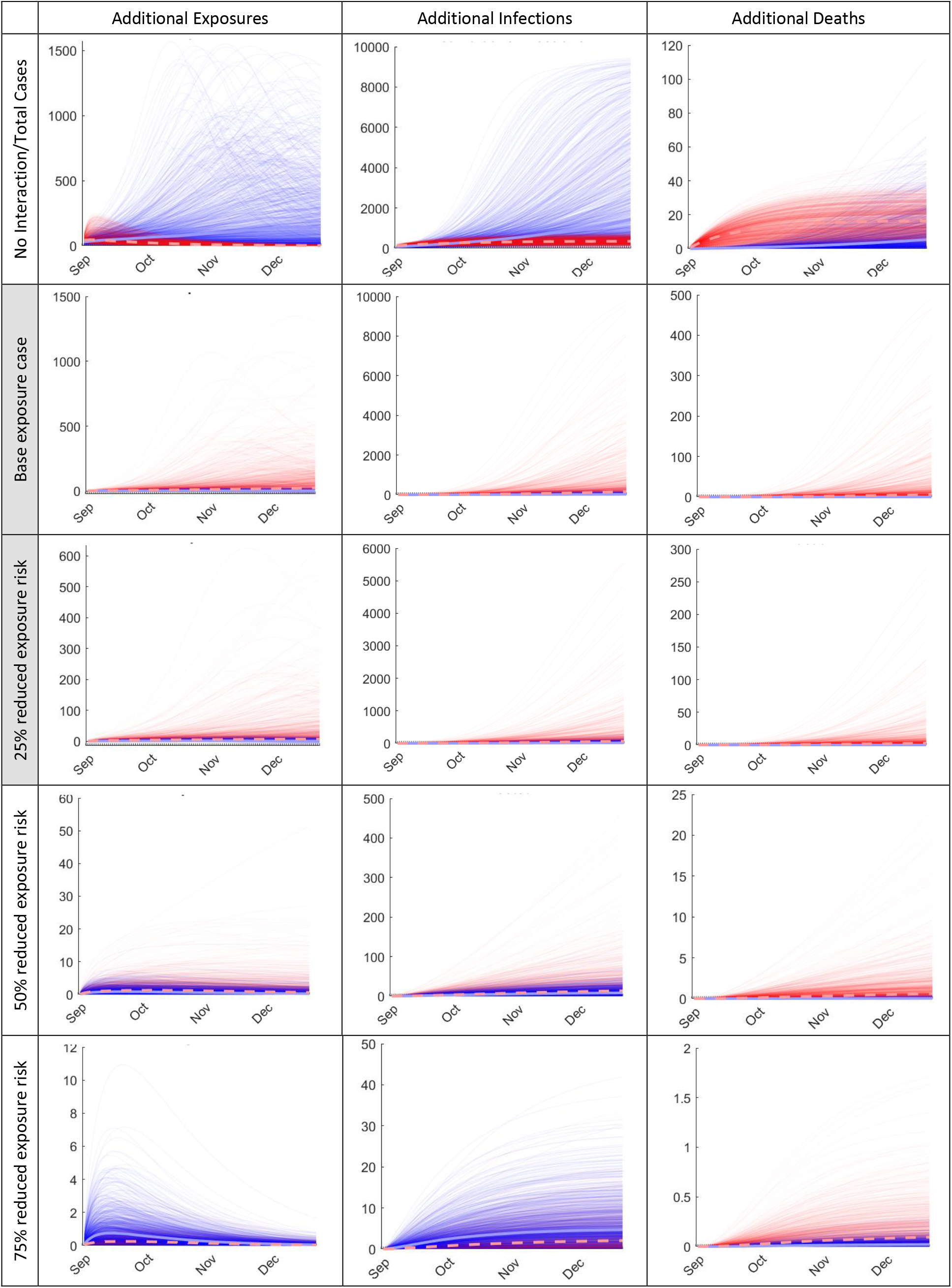
Additional (red) community and (blue) campus exposures, infections, and mortality due to community-x-campus cross-exposure (prevalence among arriving students varied between 0.1% to 2%). No interaction: <u>total</u> outcomes assuming no interaction between school and community). Base case: <u>additional</u> outcomes due to campus reopening assuming little-to-no campus semester operation precautions, compliance, or effectiveness. Shaded rows (most likely cases): <u>additional</u> outcomes assuming likely and ideal cases for campus operation precautions, adherence, and effectiveness. Bottom row: <u>additional</u> outcomes under best case scenario assuming very high campus semester operation precautions, compliance, and effectiveness.

The two more realistic scenarios result in a total of (for 25% exposure reduction) 1 to 5,577 additional community infections (median: 56, SD: 516) and 0 to 272 additional community deaths (median: 3, SD: 24), and (for 50% exposure reduction) 0 to 464 additional community infections (median: 14, SD: 45) and 0 to 23 additional community deaths (median: 1, SD: 2). For completeness, the idealistic best-case scenario results in 0 to 33 additional community infections (median: 2, SD: 4) and 0 to 2 additional community deaths (median: 0, SD: 0.2).

**Table 4.** Predicted monthly total community outcomes (per 10,000) assuming (a) no university interaction, (b) additional community resident outcomes, and (c) additional university student outcomes. Expo: exposures; Infect: infections; Mort: mortality. Tabulated values are medians; parentheses indicate 95% probability ranges given the noted uncertainties in many assumptions and inputs.

| Scenario<br>(Exposure<br>Risk) | September |  |  | October |  |  | November |  |  | December |  |  |
| --- | --- | --- | --- | --- | --- | --- | --- | --- | --- | --- | --- | --- |
|  | Expo | Infect | Mort | Expo | Infect | Mort | Expo | Infect | Mort | Expo | Infect | Mort |
| **a. Community Outcomes Assuming no University Interaction (per 10,000)** |  |  |  |  |  |  |  |  |  |  |  |  |
| Monthly<br>Totals | 11<br>(0, 64) | 158<br>(122, 362) | 6<br>(4, 18) | 4<br>(0, 30) | 175<br>(122, 455) | 7<br>(4, 26) | 1<br>(0, 14) | 181<br>(122, 486) | 7<br>(4, 28) | 1<br>(0, 8) | 182<br>(122, 495) | 7<br>(4, 29) |
| **b. Additional Community Outcomes Due to Reopening (total)** |  |  |  |  |  |  |  |  |  |  |  |  |
| Base case | 6<br>(1, 24) | 9<br>(2, 26) | 0<br>(0, 1) | 11<br>(1, 69) | 32<br>(6, 126) | 1<br>(0, 4) | 13<br>(1, 148) | 78<br>(10, 415) | 3<br>(0, 10) | 14<br>(1, 180) | 124<br>(14, 755) | 5<br>(1, 19) |
| 25%<br>reduced<br>exposure | 3<br>(1, 14) | 6<br>(2, 17) | 0<br>(0, 1) | 4<br>(0, 23) | 17<br>(4, 56) | 1<br>(0, 2) | 6<br>(0, 42) | 32<br>(5, 147) | 1<br>(0, 5) | 6<br>(0, 57) | 48<br>(7, 263) | 2<br>(0, 7) |
| 50%<br>reduced<br>exposure | 1<br>(0, 4) | 3<br>(1, 7) | 0<br>(0, 0) | 1<br>(0, 3) | 6<br>(1, 16) | 0<br>(0, 1) | 1<br>(0, 3) | 9<br>(2, 23) | 0<br>(0, 1) | 0<br>(0, 2) | 10<br>(2, 29) | 1<br>(0, 1) |
| 75%<br>reduced<br>exposure | 0<br>(0, 1) | 1<br>(0, 2) | 0<br>(0, 0) | 0<br>(0, 0) | 1<br>(0, 3) | 0<br>(0, 0) | 0<br>(0, 0) | 1<br>(0, 4) | 0<br>(0, 0) | 0<br>(0, 0) | 1<br>(0, 4) | 0<br>(0, 0) |
| **c. Additional University Outcomes Due to Reopening (total)** |  |  |  |  |  |  |  |  |  |  |  |  |
| Base case | 0<br>(0, 5) | 1<br>(0, 10) | 0<br>(0, 0) | 1<br>(0, 8) | 3<br>(0, 30) | 0<br>(0, 0) | 0<br>(0, 10) | 5<br>(0, 56) | 0<br>(0, 0) | 0<br>(0, 8) | 6<br>(0, 76) | 0<br>(0, 0) |
| 25%<br>reduced<br>exposure | 0<br>(0, 4) | 1<br>(0, 9) | 0<br>(0, 0) | 0<br>(0, 5) | 2<br>(0, 22) | 0<br>(0, 0) | 1<br>(0, 5) | 3<br>(0, 36) | 0<br>(0, 0) | 0<br>(0, 5) | 4<br>(0, 46) | 0<br>(0, 0) |
| 50%<br>reduced<br>exposure | 0<br>(0, 3) | 1<br>(0, 7) | 0<br>(0, 0) | 0<br>(0, 2) | 1<br>(0, 14) | 0<br>(0, 0) | 0<br>(0, 1) | 1<br>(0, 19) | 0<br>(0, 0) | 0<br>(0, 1) | 2<br>(0, 21) | 0<br>(0, 0) |
| 75%<br>reduced<br>exposure | 0<br>(0, 2) | 1<br>(0, 6) | 0<br>(0, 0) | 0<br>(0, 1) | 1<br>(0, 11) | 0<br>(0, 0) | 0<br>(0, 0) | 1<br>(0, 13) | 0<br>(0, 0) | 0<br>(0, 0) | 1<br>(0, 13) | 0<br>(0, 0) |

The corresponding impact of the community on student outcomes ranges from, for the base case, in a total of 0 to 390 additional student infections (median: 21, SD: 38) and 0 to 2 additional student deaths (median: 0, SD: 0.2) to 0 to 42 additional student infections (median: 5, SD: 6) and 0 additional student deaths (median: 0, SD: 0.04) for the idealistic case (75% exposure reduction). The two more likely cases result in (for 25% exposure reduction) 0 to 279 additional student infections (median: 17, SD: 30) and 0 to 1 additional student deaths (median: 0, SD: 0.1), and (for 50% exposure reduction) 0 to 115 additional student infections (median: 8, SD: 12) and 0 to 1 additional student deaths (median: 0, SD: 0.06).

To estimate the impact of school size and location (urban, rural), Figure 6 compares results under other student-to-community population sizes, assuming the same realistic arrival prevalence and campus operation precautions, compliance, and effectiveness scenarios. While differences intuitively exist in raw totals, results are similar and scale-invariant after adjusted for population size. For example, multiplying results by 2.5 for the second case of 40,000 residents produces similar curves to those for the first case of 100,000 residents. Insights from most above results therefore can be viewed as generalizable to many other settings until further analyses can be conducted for any specific university and city size.

**Figure 6.**
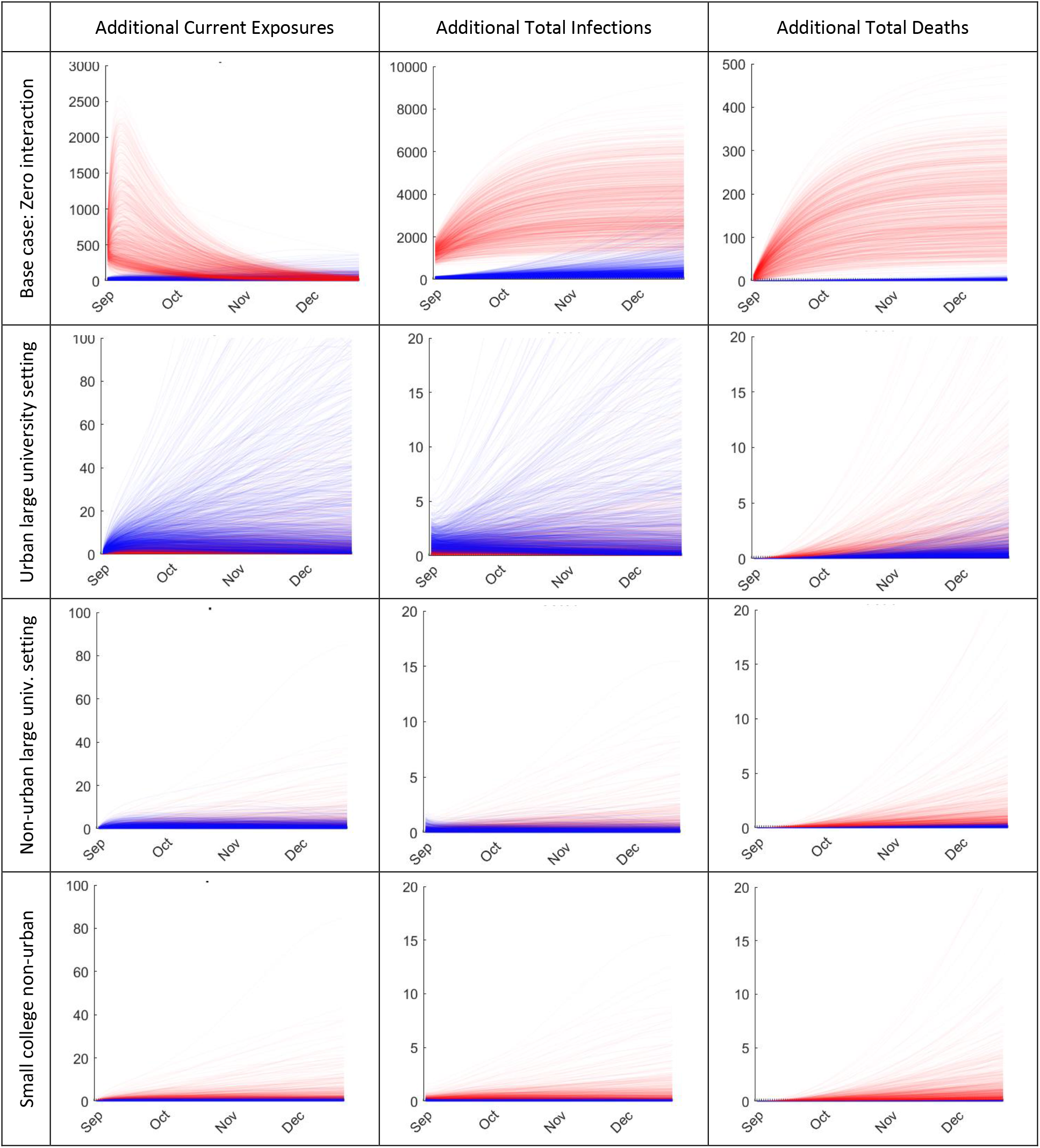
Impact of school-to-community population sizes on predicted additional community resident (red) and student (blue) current exposures, total infections, and total deaths per 10,000 individuals, assuming 1% prevalence among returning students and effective campus operations precautions (50% *R_0_* reduction). Urban large university: 10,000 students, 100,000 community residents; Non-urban large university: 10,000 students, 40,000 community residents; Small college non-urban: 2,000 students, 40,000 community residents.

Finally, factorial sensitivity analysis produced the relative parameter rankings shown in Table 2, which follow intuition and serve as further model validation. The most important statistically significant factors (main effects) affecting total campus infections were recovery time, *R_0,campus_*, incubation time, initial prevalence among arriving students, and *R_0,community_*. Similarly for total community infections, the most significant factors were recovery time, *R_0,community_*, incubation time, initial prevalence among community residents, *R_0,campus_*, *p_j_*, infection-to-death duration, and campus-community cross-exposure. Numerous interaction terms also were significant in both cases, as would be expected in such a model. Similar results for mortality are summarized in the right-most column of Table 2.

## Discussion

The Covid-19 pandemic continues to be a significant public health crisis, with infections and mortality in many regions meeting or exceeding those in early 2020 before physical distancing and work/school closures were implemented. With many colleges and universities reconsidering opening plans over the next several semesters, model-based analyses can help inform these important decisions. The large number of institutions that already reversed fall 2020 decisions also underscores this importance.

Perhaps the most important insight from results is that predicted infections and mortality from campus reopening are highly variable due to a combination of random chance, input, scenario, effectiveness, and compliance assumptions. Predicting results with any certainty is nearly impossible, despite some assurances otherwise. While regular (pre-Covid-19) campus operations could be disastrous, community and student harm even under best-case scenarios still could occur. Several other important implications exist.

First, decisions whether to open in future academic terms should include updated model inputs, understanding of dynamics, and projections of local conditions. While conditions may exist under which reopening is relatively safe, at the present time these appear in the minority. Since any trajectory within current model intervals could occur by definition, reopening decisions also should consider these ranges rather (than averages alone, which are less relevant without their probabilistic context).

Second, clear criteria should be established under which a campus should tighten policies or close altogether, including contingency plans and methods for rapidly detecting when such conditions are approaching. Absent these *a priori*, any campus reopening might be considered a breach of public trust if not negligent. Since it appears probable that at some point during the fall 2020 semester many schools may need to transition to fully virtual operations, families and public health officials should plan accordingly. A related concern is infections that might be caused elsewhere by students traveling home after the SARS-CoV-2 virus has spread among them, underscoring the importance of rapid trend detection and action.

Third, contact tracing infrastructure and effectiveness should be examined and aggressively addressed if needed. In many scenarios, large numbers of potential exposures may occur weekly, possibly exceeding some university timely tracing capabilities. While many schools report creating impressive test capacities, whether sufficient capacity exists then to trace contacts is unclear – presupposing detected positive students can (or are willing to) identify most contacts including community interactions. Revisiting whether sufficient isolated living space exists for detected positive students, based on predicted weekly infections, also may be important.

Finally, given likely similar uncertainties at least through the next few academic terms, viable alternatives to 16-week on-campus semesters might be further developed, as some have proposed. In other socioeconomic sectors, most notably healthcare, Covid-19 disruptions resulted in creation and refinement of significantly new ways to meet needs, some being improvements that continue to evolve over time. Strong and similar motivations may exist to re-invent a large and important sector of society.

Like any model-based analysis, results herein have some limitations and simplifications. A common barrier in such models is data availability for input estimation and results validation (hence our search-based approach). The deterministic ODE modeling framework ignores inherent variability and population heterogeneity,^116^ motivating our use of Monte Carlo analysis, parameter search repetitions, and varied scenarios. Standard model simplifications include limiting the number of populations (e.g., one overall homogenous community or student population), limiting spread to just SARS-CoV-2 (e.g., ignoring seasonal influenza, the substance abuse co-epidemic, ^75,76^ and co-spread impacts), and not time-varying precaution compliance as concerns and vigilance relax or heighten over time. Some scenarios also were included for potential insights rather than being feasible in practice (e.g., 75% reduction in *R*_0_, near 100% precaution compliance). Results nonetheless offer valuable insights into the range of potential community and student impacts, intervention effectiveness, and inflection criteria at which, if approaching, campuses should close.

Further work could expand on these results, including addressing some of the above simplifications made in the interest of time. Work also could occur to determine combined conditions (reduced prevalence, vaccine effectiveness, improved precaution methods, etc.) under which outcomes are both safer and more certain. City- or state-wide decision-making also might be considered in the future, such as coordinating across schools to alternate on-campus periods or limiting combined student densities to reduce net community impacts.

## Conclusion

Controlling the Covid-19 pandemic over the next several semesters is extremely critical. Computer models can offer valuable insights to important decisions, including potential community and campus impacts from university reopening. The analysis summarized herein underscores three key points: (1) exact outcomes over a 16-week semester can differ significantly under different assumptions and by chance alone, (2) off-campus impacts on local community residents could be significant, and (3) even under the best assumptions outcomes appear fairly uncertain but with near certainty non-zero. Similar analyses thus might investigate alternate approaches to provide valuable university education models until spread is better controlled.

## Data Availability

No data were used in this study. Source computer code is available from the authors.

## Acknowledgements

This research was supported in part by the National Science Foundation (CMMI-1742521) and National Institute of Drug Abuse (R21DA046776-01). The content is solely the responsibility of the authors and does not represent official views of either funder. The authors thank Basma Bargal for help with references.

